# Consuming less ultra-processed food is associated with inadequate protein intake among vegan dieters

**DOI:** 10.1101/2023.12.11.23299823

**Authors:** Alice Erwig Leitão, Gabriel P. Esteves, Bruna Caruso Mazzolani, Fabiana Infante Smaira, Martin Hindermann Santini, Heloísa C. Santo André, Bruno Gualano, Hamilton Roschel

## Abstract

**Importance:** Major concerns regarding vegan dieters are whether they meet protein and essential amino acids (EAA) recommendations, and how reliant they are on ultra-processed foods (UPF).

**Objectives:** To investigate whether vegan dieters meet protein and EAA recommendations. As secondary objectives, to determine UPF intake and potential predictors of inadequate protein intake in this population.

**Design:** A survey conducted between September 2021 and January 2023.

**Setting:** Brazil.

**Participants:** Vegan dieters of both sexes, aged 18 years or older, following a vegan diet for at least 6 months.

**Exposure:** Adherence to a vegan diet, and unprocessed and minimally processed foods (UMPF) and UPF consumption.

**Main outcome measures:** Protein and EAA intake, and food consumption according to processing level (Nova classification).

**Results:** One thousand and fourteen participants completed the survey, and 774 confirmed vegan dieters with adequate food recalls were included in the analysis. Most participants (74%) met daily protein intake according to the Recommended Dietary Allowance (RDA) (median: 1.12 g·kg^−1^·day^−1^, 95%CI 1.05; 1.16). Median EAA intake was also above RDA (with 71–91% meeting recommendations). Median UMPF intake was 66.5% (95%CI 65.0; 67.9) of total energy intake (TEI), whereas UPF consumption was 13.2% TEI (95%CI 12.4; 14.4). Adjusted logistic regression models showed that consuming protein supplements or textured soy protein, higher caloric, and higher UPF intakes were associated with reduced odds of inadequate protein intake, and that higher UMPF intakes were associated with increased odds of inadequate protein intake.

**Conclusions and Relevance:** The majority of vegan dieters attained protein and EAA intake recommendations, largely based their diet on UMPF, and had a significantly lower proportion of UPF as compared to previous reports on vegans and overall Brazilian population. Importantly, participants consuming less UPF more likely exhibited inadequate protein intake, suggesting the importance of ultra-processed proteins for this population.

**Key points:** *Question:* Do vegan dieters meet protein needs, and how reliant are they on ultra-processed foods (UPF)?

*Findings:* In this large survey including 774 vegan dieters, 74% had adequate daily protein intake. Unprocessed and minimally processed foods and UPF consumption accounted for 66.5% and 13.2% of total energy intake. Consuming protein supplements and textured soy protein, and higher caloric and UPF intakes were associated with reduced odds of inadequate protein intake, whereas higher UMPF intakes were associated with increased odds of inadequate protein intake.

*Meaning:* Most vegan dieters attain protein recommendations, while being less likely to do so when consuming less UPF.

## INTRODUCTION

Veganism is a worldwide growing lifestyle that supports abstaining from the use of animal products, leading to a dietary pattern that excludes meat, fish, poultry, dairy, eggs, and honey, among other animal-derived foods (henceforth referred to as ‘vegan diet’).^1,2^

Refraining from protein-rich animal foods has generated ongoing controversy as to whether vegan dieters can adequately meet protein requirements.^3^ Although studies show they can have borderline adequate protein intake – albeit slightly reduced *vs.* omnivorous^4^ – it is unknown whether protein sources habitually consumed by vegans allow for adequate essential amino acids (EAA) intake.

A range of plant-based meat and dairy substitutes (PMDS) have been formulated with the claim of being practical, protein-rich complements to a vegan diet.

Overconsumption of these products may, however, disrupt the alimentary basis of vegan diets – unprocessed and minimally processed (UMPF) plant-based foods, such as fruits, vegetables, and whole grains – and lead to an increase in ultra-processed foods (UPF) intake.^5,6^ The effects of UPF intake on overall health have been widely debated,^7–9^ warranting the investigation of food consumption as a function of processing level in vegan population. Therefore, the primary aim of this study was to describe protein and EAA intake in a large cohort of individuals that follow a vegan diet, with a secondary aim of assessing food intake by processing level, according to the Nova classification system, and investigating potential predictors of inadequate protein intake in this cohort.

## METHODS

### Study design and participants

Data herein derive from The Vegan Eating Habits and Nutritional Evaluation Survey (VEGAN-EatS), a cross-sectional survey conducted between September 2021 and January 2023. We have previously published an analysis of disordered eating attitudes and food choice motives on this population stemming from this survey.^10^ The current study expands on the topic by investigating the dietary profile of vegan diets, with the topical issue of protein, EAA intake, and food processing in this population.

Participants were recruited through advertisements on social media platforms and included individuals of both sexes, aged ≥18 years, currently living in Brazil, with ability to read and internet access. Participants completed an online survey using Google Forms platform (Google® LLC).

This study was approved by the local ethical committee (CAAE: 77624517.8.0000.5357) and conducted according to the Helsinki declaration. All participants signed a digital Informed Consent Form. This manuscript adheres to the STROBE-nut reporting guidelines.^11^

### Evaluation tool

The survey included questions regarding participants general characteristics, self-reported anthropometric data, and the vegan lifestyle. Macro and micronutrient, amino acids, and food intake according to processing level were assessed by food diary. Participants received an instructional video on how to fulfil the diary and fully report quantity and type of foods and beverages consumed within the previous habitual 24 hours, including a precise level of detail on food preparations and ingredients, allowing best practices for Nova food classification.^12,13^ Total energy (kcal), macronutrient (grams), and relative contribution of each Nova food processing category to total energy intake (% TEI) were calculated. We also calculated the relative contribution stemming from proteins according to food processing. Details on foods classification according to Nova are in eTable 1 in Supplement 1.

Food diaries were quantified with a specific software (Nutritionist Pro version 7.3, Axxya Systems) utilizing the USDA database. When nutritional information was not available, we searched the literature or directly contacted food companies. Amino acid composition was available for most (94.2%) of the consumed foods.

### Missing data

Due to missingness in body weight and height (27.9%), individuals with and without these data were compared and analyzed for potential associations between missingness and variables (eTable 2 in Supplement 1). There were only subtle differences between subgroups across all variables. As protein and amino acid intake adequacy requires assessing intake relative to body weight, observations with missing variables were dropped (n=216) and complete cases (n=554) were used for the main analysis. We also provided a complementary analysis in which missing body weight was imputed through multiple imputation,^14^ using the “mice” package^15^ in R set to 5 iterations and the classification and regression trees method. A matrix of predictors of body weight within the dataset were selected (age, sex, income, exercise habits, motivation to shift to a vegan diet, total energy intake and protein intake). Averaged body weight value from these 5 imputed datasets was used for descriptive statistics and model adjustment.

### Statistical analysis

Descriptive data are presented as median and interquartile range (IQR) for continuous and as absolute and relative frequency (n[%]) for categorical variables. 95% confidence intervals (95%CI) were calculated surrounding the median using bootstrap method through the “boot” package in R, set to 10,000 iterations.

Protein and amino acids intake (mg·kg^−1^·d^−1^) were compared to the Recommended Dietary Allowances (RDA) from the Dietary Reference Intakes (DRIs).^16^

Median protein and amino acid contribution of individual food items was calculated, and the top 30 or 10 food items for protein and EAA, respectively, were plotted. To further assess potential determinants of inadequate protein intake, participants were classified as having inadequate (<0.8 g·kg^−1^) or adequate (≥0.8 g·kg^−1^) protein intake,^16^ and logistic regression models were used considering protein intake status as the outcome variable. Protein supplements (Yes/No) or texturized soy protein (Yes/No) consumption were used as binary predictors. Quartiles were calculated for continuous variables (*e.g.,* kilocalories, UMPF, UPF, unprocessed, and ultra-processed protein intake) and utilized as categorical predictors. Models were adjusted for age, sex, and body weight. Alpha level was set at 0.05.

All data cleaning, exploration, and visualizations were performed using R (Version 4.2.2) and RStudio (Posit Software, PBC), with the *dplyr* and *ggplot2* packages.

## RESULTS

One thousand and fourteen participants completed the survey. During nutritional analyses, 43 were excluded for not fully adhering to a vegan diet, and 197 due to insufficient report on foods and portions. Data on 774 confirmed vegan dieters was available. Since 216 participants did not report body weight, relative protein and amino acid intakes were available for 558 individuals (eFigure 1 in Supplement 1 for study flowchart).

Median age and BMI were 29 (IQR 24; 35) years and 22.6 (IQR 20.3; 24.8) kg·m^2 −1^. Most were female (82%), had a high educational level (*i.e.,* postgraduate) (36%), within socioeconomic class B (38%), reported neither alcohol consumption (42%) nor smoking habits (91.2%), exercised from 3-6 hours/week (23%), and adhered to a vegan diet for ≥5 years (28%) (Table 1). Participants had a median intake of 1.782 (1.385; 2.227) kilocalories, with 60%, 14% and 26% distribution of carbohydrate, protein, and fat, respectively. Median protein intake was 1.12 g·kg^−1^·day^−1^ (95%CI 1.05; 1.16, Figure 1, panel A), whereas dietary fiber intake was 44 g·day^−1^ (IQR 31; 61) (Table 1). Other micronutrients can be found in eTable 3 in Supplement 1.

**Figure 1.**
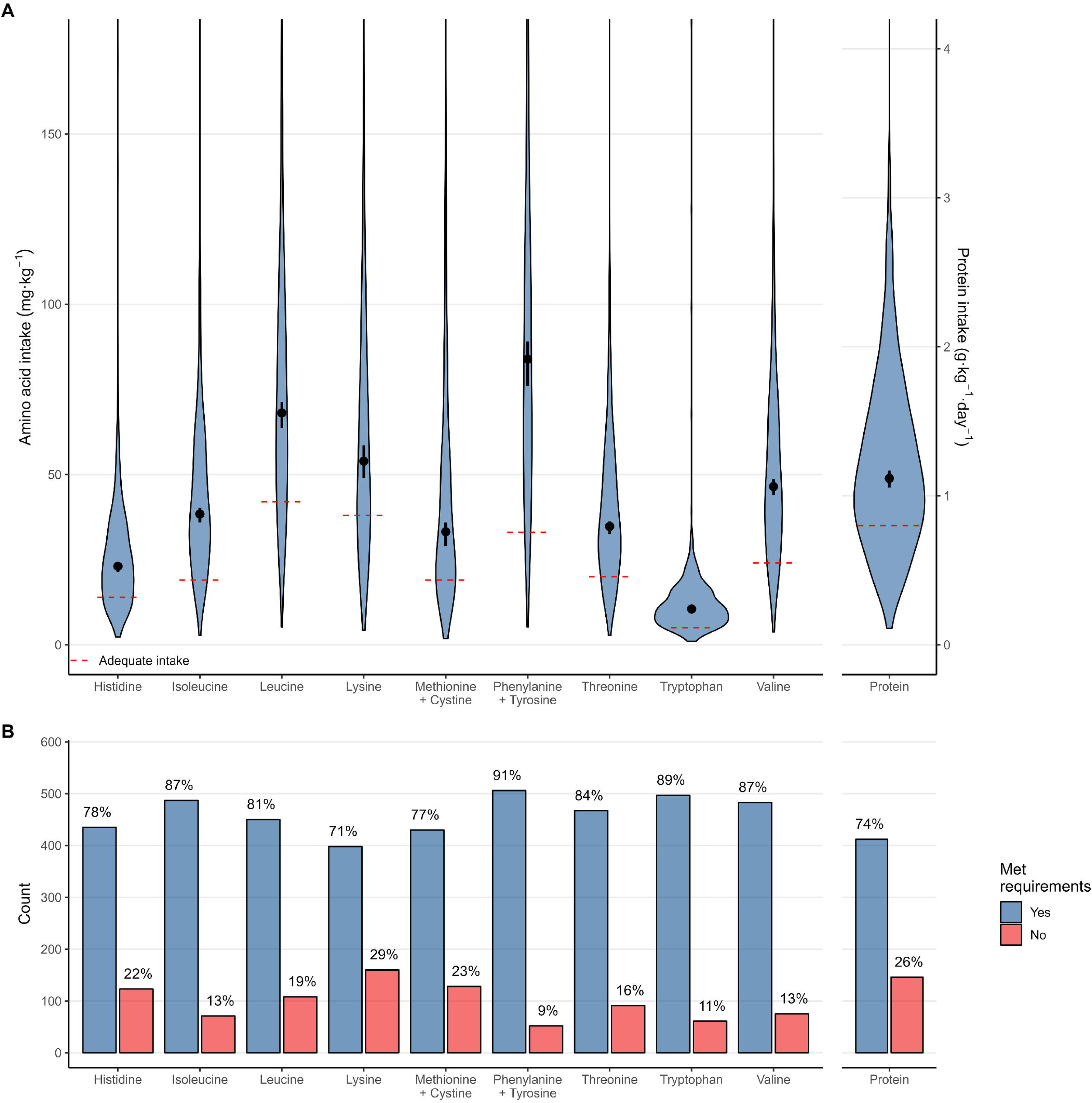
Essential amino acids intake and proportion of individuals meeting RDAs. Caption: Panel A: Violin plots showing the distribution of essential amino acids intake relative to body mass, with the red dashed line indicating the respective RDA and dots showing median values and 95% confidence intervals. Panel B: Count and proportion of individuals meeting RDA for essential amino acids intake.

**Table 1.** Sample characteristics.

| Characteristic | Overall, N = 774 | Female, N = 637 | Male, N = 137 |
| --- | --- | --- | --- |
| **Age** | 29 (24, 35) | 28 (24, 35) | 31 (25, 35) |
| **Body weight (kg)** | 60 (54, 71) | 59 (53, 66) | 74 (65, 82) |
| **Height (cm)** | 165 (160, 170) | 163 (159, 168) | 175 (170, 180) |
| **BMI (g/kg<sup>2</sup>)** | 22.6 (20.3, 24.8) | 22.1 (20.1, 24.3) | 23.9 (22.7, 25.9) |
| **Educational level** |  |  |  |
| College education or technician, complete | 242 (31%) | 190 (30%) | 52 (38%) |
| Elementary school, completed | 2 (0.3%) | 1 (0.2%) | 1 (0.7%) |
| Elementary school, incomplete | 2 (0.3%) | 2 (0.3%) | 0 (0%) |
| High school, completed | 55 (7.1%) | 50 (7.8%) | 5 (3.6%) |
| High school, incomplete | 18 (2.3%) | 18 (2.8%) | 0 (0%) |
| Postgraduate | 280 (36%) | 240 (38%) | 40 (29%) |
| Undergoing college or technician education | 175 (23%) | 136 (21%) | 39 (28%) |
| **Income** |  |  |  |
| A class | 41 (5.3%) | 29 (4.6%) | 12 (8.8%) |
| B class | 296 (38%) | 243 (38%) | 53 (39%) |
| C class | 196 (25%) | 161 (25%) | 35 (26%) |
| D/E class | 241 (31%) | 204 (32%) | 37 (27%) |
| **Smoking status (smoker)** | 68 (8.8%) | 53 (8.3%) | 15 (11%) |
| **Alcohol consumption** |  |  |  |
| No alcohol consumption | 324 (42%) | 267 (42%) | 57 (42%) |
| Once to twice a month | 147 (19%) | 127 (20%) | 20 (15%) |
| Twice to four times a month | 222 (29%) | 176 (28%) | 46 (34%) |
| Twice to three times per week | 72 (9.3%) | 60 (9.4%) | 12 (8.8%) |
| Four or more times per week | 9 (1.2%) | 7 (1.1%) | 2 (1.5%) |
| **Habitual physical exercise** |  |  |  |
| Does not exercise | 142 (18%) | 126 (20%) | 16 (12%) |
| 1-2 hour/week | 156 (20%) | 136 (21%) | 20 (15%) |
| 3-4 hours/week | 176 (23%) | 145 (23%) | 31 (23%) |
| 5-6 hours/week | 176 (23%) | 142 (22%) | 34 (25%) |
| 7 hours/week or more | 124 (16%) | 88 (14%) | 36 (26%) |
| **How long as a vegan** |  |  |  |
| Less than one year | 110 (14%) | 89 (14%) | 21 (15%) |
| 1 to 2 years | 174 (22%) | 141 (22%) | 33 (24%) |
| 2 to 3 years | 153 (20%) | 127 (20%) | 26 (19%) |
| 3 to 4 years | 118 (15%) | 102 (16%) | 16 (12%) |
| 5 or more years | 219 (28%) | 178 (28%) | 41 (30%) |
| Supplement use | 592 (76%) | 484 (76%) | 108 (79%) |
| **Nutritional intake** |  |  |  |
| Kilocalories (kcal) | 1,782 (1,385, 2,227) | 1,725 (1,365, 2,127) | 2,200 (1,729, 2,914) |
| Protein (g) | 70 (48, 94) | 67 (48, 89) | 86 (60, 126) |
| Carbohydrate (g) | 268 (204, 346) | 255 (198, 325) | 342 (261, 439) |
| Fat (g) | 53 (37, 72) | 51 (36, 67) | 62 (44, 88) |
| Protein (g·kg<sup>-1</sup>) * | 1.12 (0.79, 1.53) | 1.11 (0.79, 1.48) | 1.14 (0.78, 1.69) |
| Carbohydrate (% TEI) | 60 (52, 65) | 60 (52, 65) | 60 (55, 65) |
| Protein (% TEI) | 14 (12, 18) | 15 (12, 18) | 15 (12, 18) |
| Fat (% TEI) | 26 (19, 31) | 25 (19, 31) | 25 (19, 29) |
| Saturated fatty acid (g) | 10 (7, 14) | 9 (6, 14) | 12 (8, 19) |
| Monounsaturated fatty acid (g) | 19 (13, 28) | 18 (12, 26) | 23 (16, 35) |
| Polyunsaturated fatty acid (g) | 16 (11, 22) | 15 (10, 21) | 20 (11, 28) |
| Trans-fatty acid (g) | 0.04 (0.01, 0.08) | 0.04 (0.01, 0.08) | 0.06 (0.02, 0.13) |
| Dietary fiber (g) | 44 (31, 61) | 41 (30, 57) | 58 (39, 76) |

Results are presented as median (interquartile range) for continuous variables and n (%) for categorical variables. BMI, body mass index. \* N = 558. Income was categorized according to the Brazilian Institute of Geography and Statistics (IBGE) in levels A ( ≥ USD 4,200), B (USD 1,350 to 4,200), C (USD 550 to 1,350), and D/E ( ≤ USD 550).<sup>50</sup>

Median intake of all EAA were significantly above RDAs (Table 2, Figure 1, panel B. Lysine had the lowest rate of adequacy (71%), with the highest being for phenylalanine and tyrosine (91%) (Figure 1, panel B). Regarding food processing, participants showed a high intake of UMPF (66.5% TEI [95%CI 65.0; 67.9]), which is significantly higher than values reported for metropolitan areas in Brazil,^17^ and low PCI (8.3% TEI [95%CI 7.6; 8.8]), PF (6.2% TEI [95%CI 5.1; 6.8]) and UPF (13.2% TEI [95%CI 12.4; 14.4]) consumption, all significantly lower than values reported for Brazilian metropolitan areas (Table 2, Figure 2).^17^ UMPF were also the main source of protein for individuals adhering to a vegan diet (61.8% [95%CI 59.4; 64.0]), followed by UPF (23.6% [95%CI 21.0; 26.3]) and PF (7.4% [95%CI 6.1; 8.7]). A sensitivity analysis considering textured soy protein as UMPF, rather than UPF, led to a drop in total caloric contribution from UPF from 13.2 to 10.7%, and from 23.6 to 14.3% in the UPF contribution to protein energy intake (eTable 4 and eFigure 2 in Supplement 1). Individual food analysis for the main sources of protein and EAA is available in eFigure 3 in Supplement 1.

**Figure 2.**
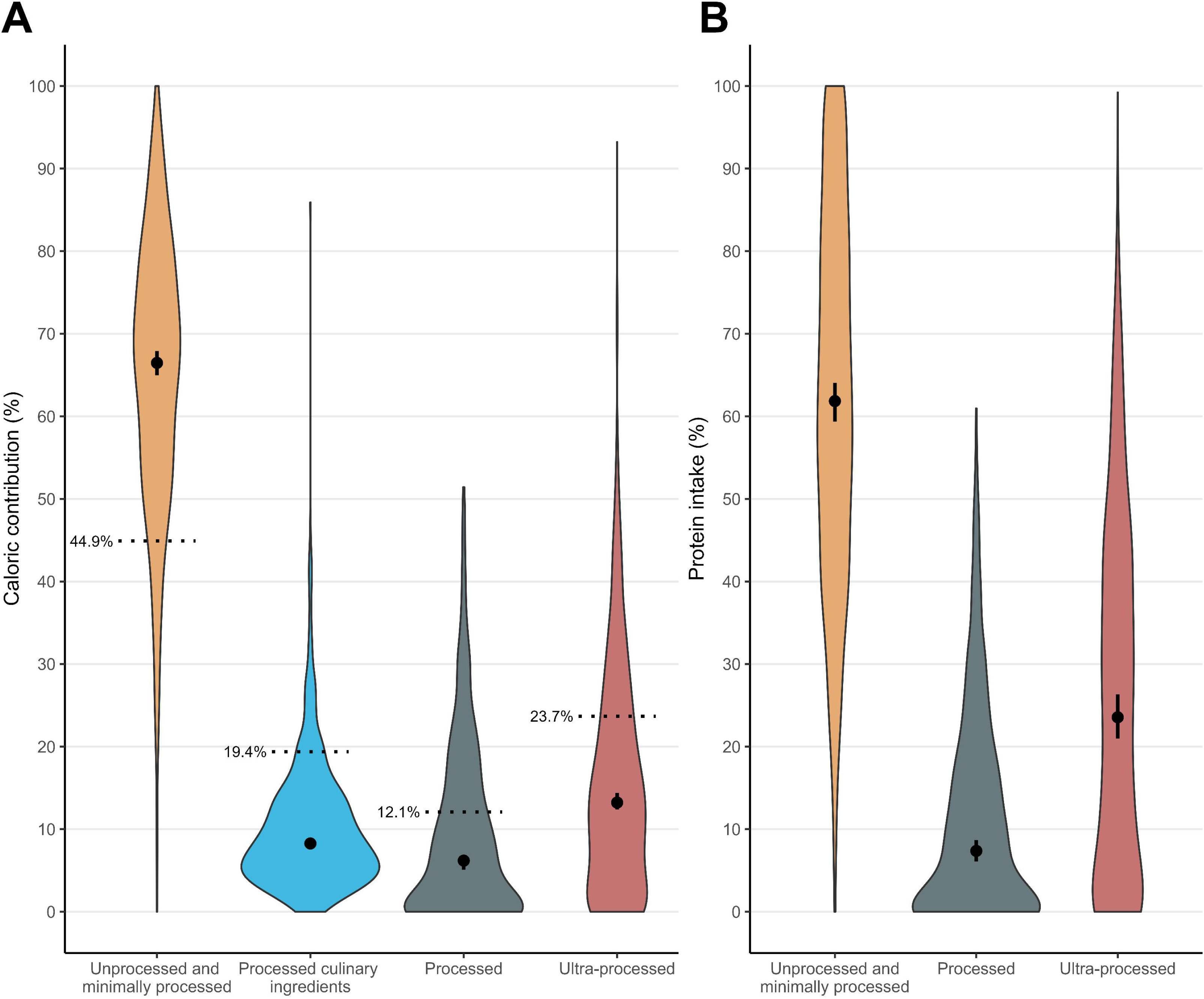
Caloric and protein intake according to Nova food processing categories. Caption: Panel A: Violin plots showing the distribution of relative caloric contribution of each food processing category. Dashed line show reference values from the Brazilian population living in metropolitan areas. Panel B: Relative contribution of each food processing category to protein intake. Dots are medians accompanied by 95% confidence intervals.

**Table 2.**
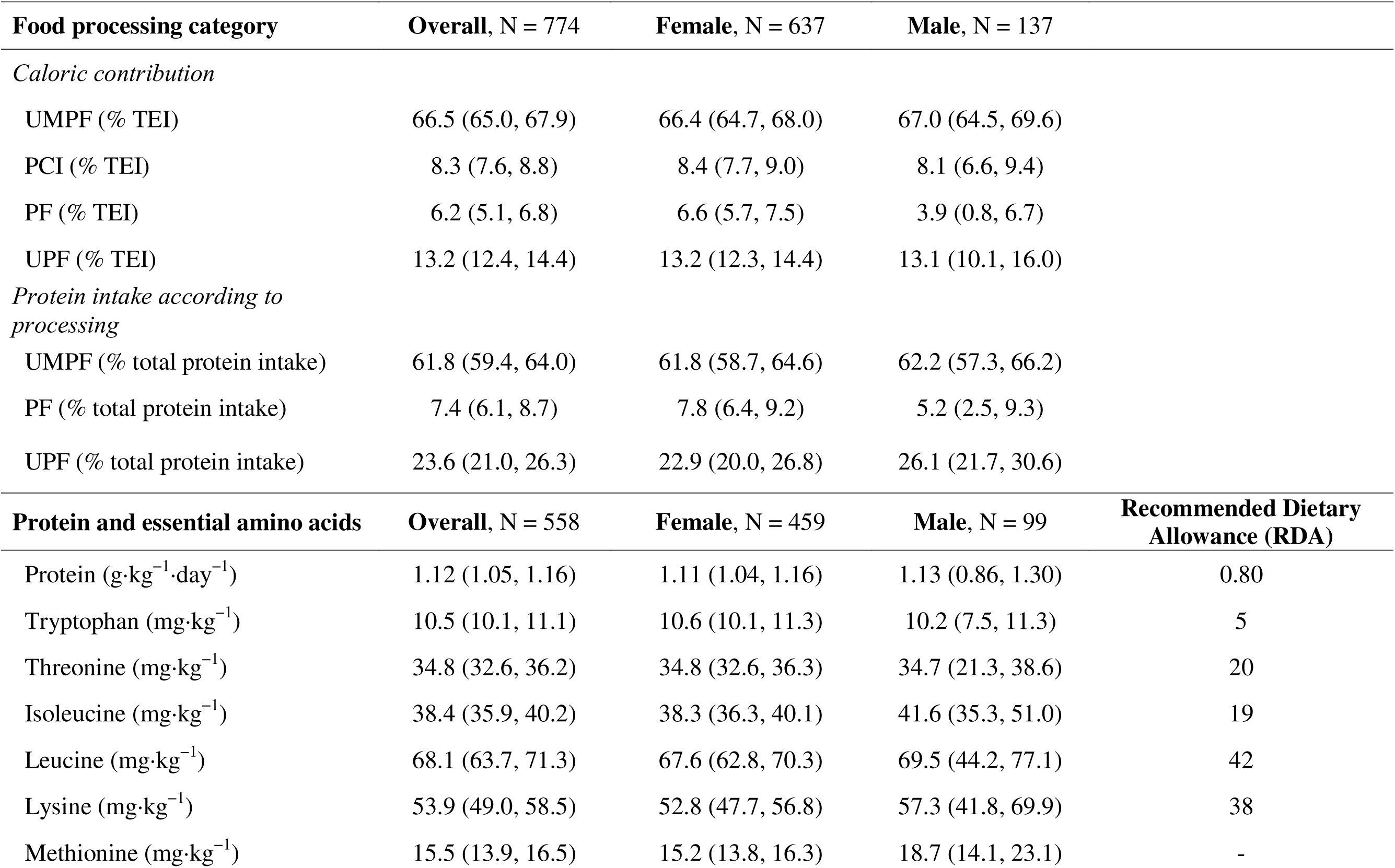
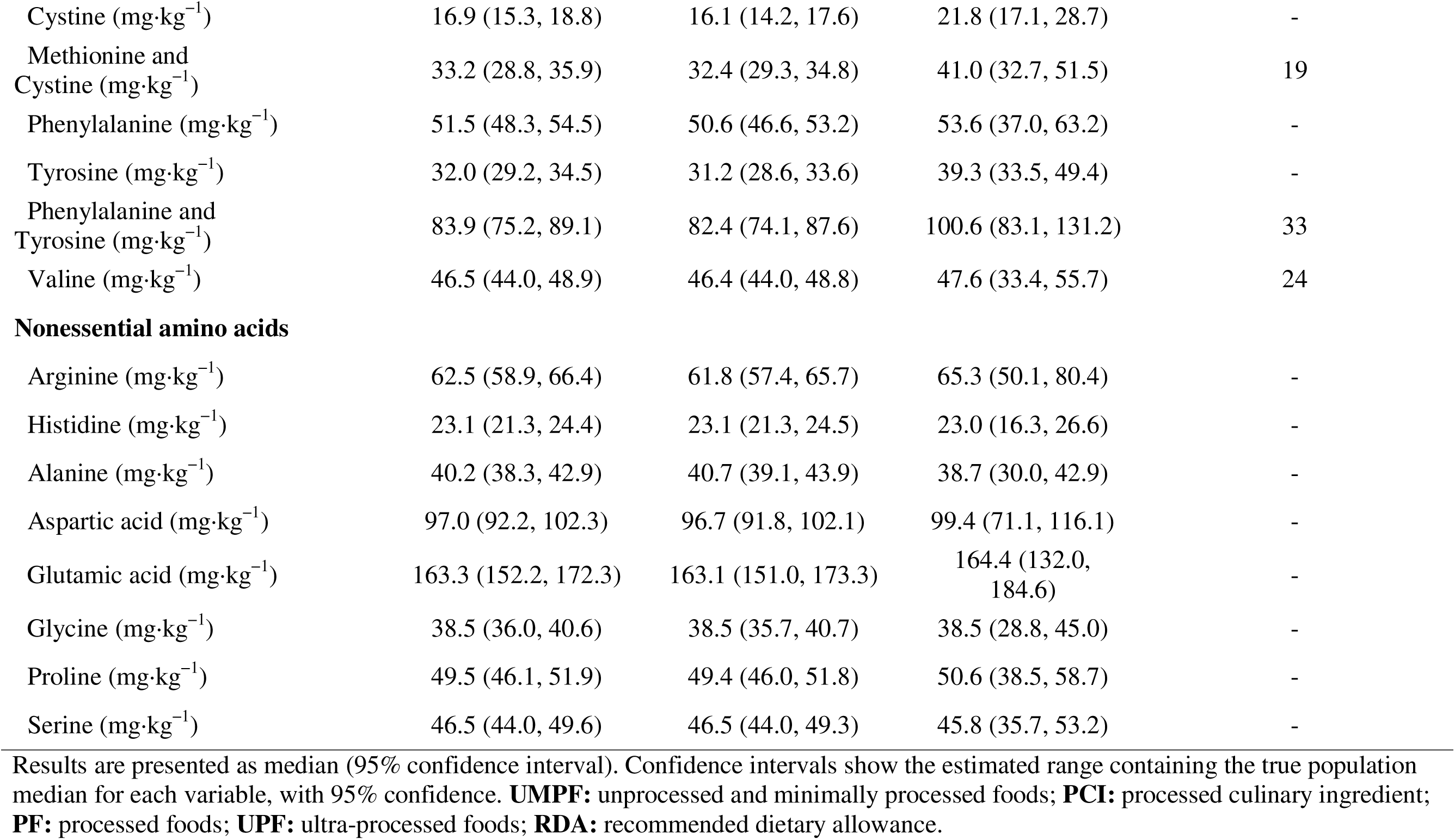
Caloric contribution and pr otein intake according to Nova food processing ca tegory and amino acid intake.

| **Food processing category** | **Overall, N = 774** | **Female, N = 637** | **Male, N = 137** |  |
| --- | --- | --- | --- | --- |
| *Caloric contribution* |  |  |  |  |
| UMPF (% TEI) | 66.5 (65.0, 67.9) | 66.4 (64.7, 68.0) | 67.0 (64.5, 69.6) |  |
| PCI (% TEI) | 8.3 (7.6, 8.8) | 8.4 (7.7, 9.0) | 8.1 (6.6, 9.4) |  |
| PF (% TEI) | 6.2 (5.1, 6.8) | 6.6 (5.7, 7.5) | 3.9 (0.8, 6.7) |  |
| UPF (% TEI) | 13.2 (12.4, 14.4) | 13.2 (12.3, 14.4) | 13.1 (10.1, 16.0) |  |
| *Protein intake according to processing* |  |  |  |  |
| UMPF (% total protein intake) | 61.8 (59.4, 64.0) | 61.8 (58.7, 64.6) | 62.2 (57.3, 66.2) |  |
| PF (% total protein intake) | 7.4 (6.1, 8.7) | 7.8 (6.4, 9.2) | 5.2 (2.5, 9.3) |  |
| UPF (% total protein intake) | 23.6 (21.0, 26.3) | 22.9 (20.0, 26.8) | 26.1 (21.7, 30.6) |  |
| **Protein and essential amino acids** | **Overall, N = 558** | **Female, N = 459** | **Male, N = 99** | **Recommended Dietary Allowance (RDA)** |
| Protein (g·kg<sup>-1</sup>·day<sup>-1</sup>) | 1.12 (1.05, 1.16) | 1.11 (1.04, 1.16) | 1.13 (0.86, 1.30) | 0.80 |
| Tryptophan (mg·kg<sup>-1</sup>) | 10.5 (10.1, 11.1) | 10.6 (10.1, 11.3) | 10.2 (7.5, 11.3) | 5 |
| Threonine (mg·kg<sup>-1</sup>) | 34.8 (32.6, 36.2) | 34.8 (32.6, 36.3) | 34.7 (21.3, 38.6) | 20 |
| Isoleucine (mg·kg<sup>-1</sup>) | 38.4 (35.9, 40.2) | 38.3 (36.3, 40.1) | 41.6 (35.3, 51.0) | 19 |
| Leucine (mg·kg<sup>-1</sup>) | 68.1 (63.7, 71.3) | 67.6 (62.8, 70.3) | 69.5 (44.2, 77.1) | 42 |
| Lysine (mg·kg<sup>-1</sup>) | 53.9 (49.0, 58.5) | 52.8 (47.7, 56.8) | 57.3 (41.8, 69.9) | 38 |
| Methionine (mg·kg<sup>-1</sup>) | 15.5 (13.9, 16.5) | 15.2 (13.8, 16.3) | 18.7 (14.1, 23.1) | - |
| Cystine (mg·kg<sup>-1</sup>) | 16.9 (15.3, 18.8) | 16.1 (14.2, 17.6) | 21.8 (17.1, 28.7) | - |
| Methionine and Cystine (mg·kg<sup>-1</sup>) | 33.2 (28.8, 35.9) | 32.4 (29.3, 34.8) | 41.0 (32.7, 51.5) | 19 |
| Phenylalanine (mg·kg<sup>-1</sup>) | 51.5 (48.3, 54.5) | 50.6 (46.6, 53.2) | 53.6 (37.0, 63.2) | - |
| Tyrosine (mg·kg<sup>-1</sup>) | 32.0 (29.2, 34.5) | 31.2 (28.6, 33.6) | 39.3 (33.5, 49.4) | - |
| Phenylalanine and Tyrosine (mg·kg<sup>-1</sup>) | 83.9 (75.2, 89.1) | 82.4 (74.1, 87.6) | 100.6 (83.1, 131.2) | 33 |
| Valine (mg·kg<sup>-1</sup>) | 46.5 (44.0, 48.9) | 46.4 (44.0, 48.8) | 47.6 (33.4, 55.7) | 24 |
| Nonessential amino acids |  |  |  |  |
| Arginine (mg·kg<sup>-1</sup>) | 62.5 (58.9, 66.4) | 61.8 (57.4, 65.7) | 65.3 (50.1, 80.4) | - |
| Histidine (mg·kg<sup>-1</sup>) | 23.1 (21.3, 24.4) | 23.1 (21.3, 24.5) | 23.0 (16.3, 26.6) | - |
| Alanine (mg·kg<sup>-1</sup>) | 40.2 (38.3, 42.9) | 40.7 (39.1, 43.9) | 38.7 (30.0, 42.9) | - |
| Aspartic acid (mg·kg<sup>-1</sup>) | 97.0 (92.2, 102.3) | 96.7 (91.8, 102.1) | 99.4 (71.1, 116.1) | - |
| Glutamic acid (mg·kg<sup>-1</sup>) | 163.3 (152.2, 172.3) | 163.1 (151.0, 173.3) | 164.4 (132.0, 184.6) | - |
| Glycine (mg·kg<sup>-1</sup>) | 38.5 (36.0, 40.6) | 38.5 (35.7, 40.7) | 38.5 (28.8, 45.0) | - |
| Proline (mg·kg<sup>-1</sup>) | 49.5 (46.1, 51.9) | 49.4 (46.0, 51.8) | 50.6 (38.5, 58.7) | - |
| Serine (mg·kg<sup>-1</sup>) | 46.5 (44.0, 49.6) | 46.5 (44.0, 49.3) | 45.8 (35.7, 53.2) | - |

Results are presented as median (95% confidence interval). Confidence intervals show the estimated range containing the true population median for each variable, with 95% confidence. **UMPF**: unprocessed and minimally processed foods; **PCI**: processed culinary ingredient; **PF**: processed foods; **UPF**: ultra-processed foods; **RDA**: recommended dietary allowance.

Adjusted logistic regression models showed that consuming protein supplements (odds ratio (OR) = 0.05 [95%CI 0.01, 0.12], p < 0.0001) and textured soy protein (OR = 0.31 [95%CI 0.16, 0.58], p < 0.001), as compared to not consuming those products, were significantly associated with reduced odds of displaying inadequate protein intake (Figure 3, panel A). The second, third and fourth quartiles of total kilocalories intake and UPF intake, and the third and fourth quartiles of ultra-processed protein intake, were all significantly associated with reduced odds of displaying inadequate protein intake (all p < 0.0001), while second, third and fourth quartiles of unprocessed protein intake was associated with increased odds of displaying inadequate protein intake (all p < 0.002) when compared to the respective first quartile (Figure 3, panel B for model coefficients, and eTable 5 in Supplement 1 for quartiles).

**Figure 3.**
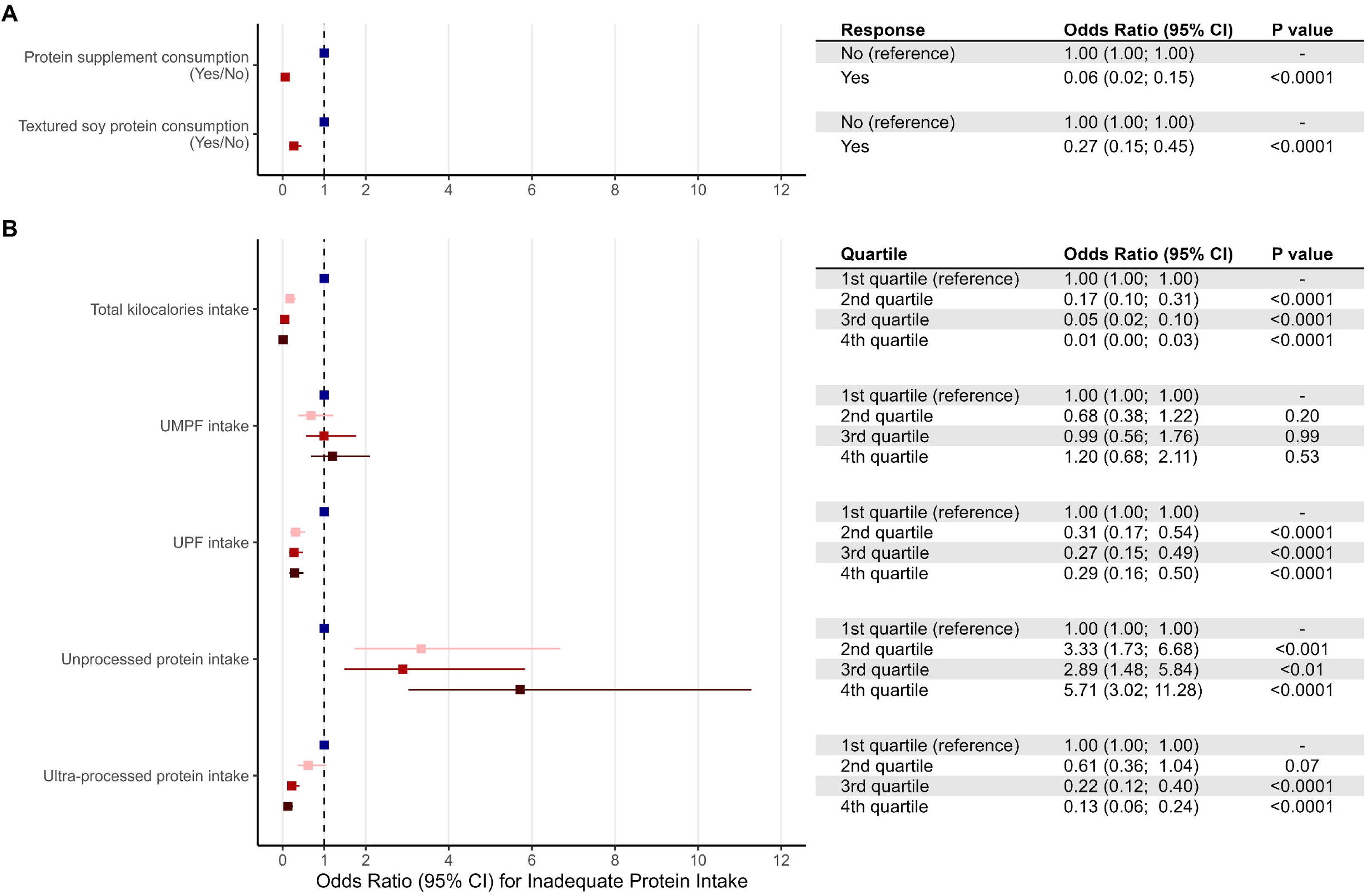
Logistic regression model results. Caption: Panel A shows the association between binary predictors (consuming protein supplements and consuming textured soy protein) and inadequate protein intake. Panel B shows association between quartiles of continuous predictors and inadequate protein intake. UMPF = unprocessed and minimally processed food; UPF = ultra-processed food. Results are presented as the odds ratio coefficients and 95% confidence intervals (CI) for having inadequate protein intake given the predictor, in comparison to the reference level.

Importantly, results remained virtually unchanged in the analyses performed on the imputed dataset (n=774), not altering the original interpretation of our results (eTable 6 and eFigure 4 in Supplement 1).

## DISCUSSION

The present study provided detailed data on protein and amino acid intake, alongside food processing level according to Nova, in a large cohort of vegan dieters. The main findings were: 1) most individuals met protein (74%) and EAA (71–91%) intake recommendations; 2) participants had a high consumption of UMPF and low consumption of PF and UPF; 3) consumption of ultra-processed protein sources were associated with decreased odds of showing inadequate protein intake, while the opposite was true for unprocessed and minimally processed protein sources.

Although previous evidence from smaller cohorts indicates that vegan diets may provide adequate protein,^4^ albeit lower then omnivorous, it was still unclear whether they deliver enough EAA. In our sample, we found that 74% of vegan dieters showed adequate protein intake, both when considering energy contribution (median 14% TEI) and daily relative intake (median 1.12 g·kg^−1^·day^−1^, 95%CI 1.05; 1.16), adhering to both Acceptable Macronutrient Distribution Range (AMDR – 10-35%) and RDA protein recommendations (0.8 g·kg^−1^·day^−1^).^16^ Importantly, most participants also met EAA recommendations, with a small proportion falling short of daily necessary amounts (9– 29%). To our best knowledge, the largest previous study to address amino acid intake in vegans was a cross-sectional analysis of the EPIC-Oxford cohort.^18^ In a relatively small sample of 98 participants, intake of amino acids was lower compared to meat-eaters. It was unclear, however, whether this difference could lead to relevant deficiencies, as authors did not compare it to recommendations.^18^ Based on a larger sample and using a higher resolution instrument, our study offers novel evidence that the majority of vegans can meet EAA recommendations.

Notwithstanding the above, a considerable part of the participants did not meet recommendations for particular EAAs. For instance, 29% and 23% fell short for lysine and the combination of methionine and cystine requirements, respectively. Indeed, the contents of these two amino acids are generally lower in plant-vs. animal-based proteins.^19,20^ Importantly, all amino acids are needed in adequate amounts to support *de novo* tissue protein synthesis,^21^ reinforcing the need for properly planned, well-balanced combinations of different plant-based protein sources to attain an optimal EAA profile. This also raises the question of whether protein requirements should be revisited when considering a plant-exclusive diet. We and others have previously demonstrated that, in vegans, when provided in adequate amounts (*i.e.,* ≥1.6 g·kg^−1^·day^−1^),^22^ plant proteins are able to fully support muscle anabolism when compared to protein-matched omnivores.^23, 24^

Assessing food intake by processing level has become an important approach from a public health perspective.^25–27^ Mounting evidence shows that increased UPF consumption favors several negative health-related outcomes, such as higher energy intake and weight gain, higher prevalence of non-transmissible chronic diseases, and overall increased mortality rate.^28–30^ Although vegan diets are traditionally based on fresh and *in natura* foods, there has been a tendency of increased UPF consumption in vegans due to proliferation of PMDS.^5^ The prevalence of vegetarianism/veganism is ever-growing, and PMDSs are an expanding market with the potential to dominate shelf spaces in supermarkets to the detriment of UMPF, possibly reducing the quality of vegan diets.^5^ A previous study in a French cohort found a 39.5% TEI of UPF consumption among vegan dieters vs. 33% TEI for meat-eaters.^6^ This confirms that the presence of UPF in vegan diets is already elevated in some regions. In our study, UMPF represented the highest caloric contribution in vegan diets (66.5% TEI), with a significantly smaller presence of UPF (13.2% TEI).

Discrepancies between the two studies are notable. Considering that both cohorts are similar in socioeconomic status and educational level, which have been demonstrated to influence consumption of UMPF,^31^ divergence in results must be explained by other reasons. Previous studies assessing food intake among the Brazilian population have shown higher UMPF and lower UPF consumption compared to other countries.^32,33^ Importantly, vegan dieters assessed in our study showed even smaller UPF consumption compared to values reported for metropolitan areas in Brazil.^17^ One may argue that, in comparison to other countries, the overall PMDS industry in Brazil may be still less active, therefore providing fewer and/or less attractive options of UPF. As there is a global trend in the protein market towards an increasing popularity of plant-based protein and a growing demand for high-quality, natural, and sustainable protein sources,^34^ one may expect that the consumption of UPF may increase among Brazilian vegan dieters in the near future as a consequence of the PMDS market expansion, warranting follow-up surveys to monitor possible impacts.

Previous studies have consistently shown associations between UPF and poor health outcomes;^9,35,36^ however, Nova UPF is a very broad category that encompasses a variety of animal-and plant-based foods, which could have contrasting impacts on human health.^37–39^ In fact, a recent study has shown that distinct subgroups of UPF are differently associated with type-2 diabetes risk, with subgroups such as fruit- and dairy-based ultra-processed desserts even showing a risk reduction.^39^ Another study found that although UPF consumption was associated with multimorbidity related to cancer and cardiometabolic diseases, this association was not seen in the subgroup of plant-based UPF,^40^ suggesting that Nova may not capture possible nuance in “quality” among different UPF. In our study, textured soy protein was an important contributor to UPF consumption. This is a challenging food item to classify, as there is significant variability in formulations considering the presence of food additives and cosmetics. This prompted us to a more conservative approach of classifying it as UPF, which is in accordance with previous research.^6^ In a sensitivity analysis considering textured soy protein as UMPF, %TEI from UPF drops from 13.2 to 10.7%. The drop in relative contribution of UPF to protein intake is even more pronounced (23.6 to 14.3%) (Table 2 and eTable 4 in Supplement 1), clearly demonstrating that reclassification of individual foods might have enormous impact on UPF contribution in vegan diets.

Indeed, we found textured soy protein to be one of the main sources of protein and EAA, followed by protein supplements, such as pea and soy protein; vegan meat substitutes (either UMPF or UPF, depending on preparation and ingredients); and a mix of vegetables and legumes, such as beans, corn, chickpeas, lentils. Collectively, these data further reinforce the strong presence of UPF protein sources in vegan diets (eFigure 3 in Supplement 1).

Adjusted logistic regressions showed that consuming protein supplements or textured soy protein, higher caloric intake, and higher UPF were associated with reduced odds of inadequate protein intake, while consuming more unprocessed protein (but not UMPF as a whole) were associated with increased odds of inadequate protein intake. These associations showed substantial magnitudes. The highest quartile of ultra-processed protein consumption associated with ∼7-fold decrease in the odds of inadequate protein intake (*vs.* the lowest quartile). Likewise, the highest quartile of unprocessed protein consumption was associated with a ∼5-fold increase in the odds for protein inadequacy. This may be partially explained by the lower energy and protein density in plant- vs. animal-derived foods,^41^ suggesting it may be challenging for vegans fully avoiding UPF to reach higher levels of protein intake without substantially increasing food (and perhaps calorie) intake.

While this does not necessarily imply that UPFs are essential for vegan dieters to meet protein recommendations, it reveals a significant reliance on UPF to attend protein requirements. Considering this, one may suggest that certain UPF, such as textured soy protein, might be recommended for this population. Despite convincing data associating the broad category of UPF with poor outcomes, it is hard to reconcile textured soy protein as having detrimental health effects, with ample evidence suggesting otherwise.^42–44^ This holds true for protein supplements, an evidence-based strategy to support muscle health^45^ also associated with protein adequacy in this study. Regarding vegan dieters, at least, unrestricted advice to avoid UPF may have unintended consequences, such as protein intake inadequacies, that warrant further investigation. This also suggests that vegans may benefit from public policies aimed to facilitate access to more natural and healthy foods, and amplify nutritional support/education for adequacy of overall food intake. Simultaneously, our data reinforce the urgent need for the development of affordable, healthier, better quality, cleaner-label and protein-rich plant-based food options by the industry.

Strengths of this study include the large sample size and use of food diaries to quantify food intake.^46^ Limitations include the cross-sectional design and self-reporting of information. Our study features a convenience sample, predominantly composed of females, with eutrophic BMI and high educational level. While this may limit generalizability of our findings, epidemiological studies confirm this as the typical sociodemographic profile of vegans.^47–49^ Nonetheless, our conclusions cannot be extrapolated to more vulnerable cohorts or different dietary patterns. Further studies are warranted to answer such questions.

In conclusion, vegan dieters mostly attained protein and EAA intake recommendations and had a significantly lower proportion of UPF as compared to previous reports on vegans and overall Brazilian population. Importantly, ultra-processed protein sources were associated with decreased likelihood of displaying inadequate protein intake, while the opposite was true for unprocessed protein sources. The role of UPF in vegan diets needs to be further investigated, as common protein sources in the vegan diet may not display the same detrimental health effects as other UPF, while contributing to protein requirements in this population.

## Author contributions

AEL, GPE, BCM, FIS and HR designed and conducted research; GPE and AEL analyzed data; and AEL, GPE, BCM, FIS, MHS, HCSA, BG and HR wrote the paper. All authors read and approved the final manuscript. The corresponding author attests that all listed authors meet authorship criteria and that no others meeting the criteria have been omitted.

## Competing interests

BG and HR have received research grants and supplement donations for scientific studies from AlzChem, Natural Alternatives International, DuPont, J.B.S., and NotCompany. BG has also received support for participation in scientific conferences, and honorarium for speaking at lectures from AlzChem. Additionally, he serves as a member of the Scientific Advisory Board for AlzChem. None of these fundings are related to the present study in any aspect.

## Funding

GPE, BM, FS, HCSA, BG and HR were supported by São Paulo Research Foundation – FAPESP (grants #2019/14820-6, #2019/14819-8, #2020/07860-9, #2022/02229-4 and #2017/13552-2). AE and MHS were supported by Coordenação de Aperfeiçoamento de Pessoal de Nível Superior - Brasil (CAPES) - Finance Code 001. HR and BG were supported by National Council for Scientific and Technological Development–CNPq (#308307/2021-6 and #301914/2017-6). These funding agencies did not have any role in any aspect of the current study.

## Data sharing statement

Data pertaining to this study can be shared, alongside statistical code, upon request.

## Data Availability

Data pertaining to this study can be shared, alongside statistical code, upon request.

## Notes

### Funding Statement

GPE, BM, FS, HCSA, BG and HR were supported by Sao Paulo Research Foundation - FAPESP (grants #2019/14820-6, #2019/14819-8, #2020/07860-9, #2022/02229-4 and #2017/13552-2). AE and MHS were supported by Coordenacao de Aperfeicoamento de Pessoal de Nivel Superior - Brasil (CAPES) - Finance Code 001. HR and BG were supported by National Council for Scientific and Technological Development-CNPq (#308307/2021-6 and #301914/2017-6). These funding agencies did not have any role in any aspect of the current study.

### Author Declarations

The protocol was approved by the ethics committee of the Hospital das Clinicas of the Faculty of Medicine of the University of Sao Paulo under the protocol CAAE: 77624517.8.0000.5357.

