## Supplement 1 for "Consuming less ultra-processed food is associated with inadequate protein intake among vegan dieters"

**eTable 1.** Examples of common vegan foods from Nutritionist Pro software and classification according to Nova system.

| **Nova food classification** | **Vegan food label in Nutritionist Pro software** |
| --- | --- |
| Unprocessed and minimally processed (UMPF) | Avocado |
|  | Chayote, Boiled with Salt, Drained |
|  | Chestnut, Dried |
|  | Chickpeas, Beans and Peas |
|  | Cocoa powder, unsweetened |
|  | Corn, Dry |
|  | Cornstarch |
|  | Cranberry, low bush or lingenberry, fresh (Alaska native) |
|  | Dates, Dry |
|  | Eggplant |
|  | Flour, All Purpose Wheat, Self-Rising, Enriched |
|  | Flour, chickpea or besan |
|  | Flour, corn, white, whole grain |
|  | Mushrooms, boiled, drained |
|  | Oat Bran, Dry |
|  | Onions, Chopped |
|  | Papaya |
|  | Parsley, Chopped |
|  | Peanut Butter, Smooth |
|  | Pearl tapioca, dry |
|  | Pepper, Black, Ground |
|  | Popcorn, popped in oil |
|  | Potatoes baked |
|  | Quinoa |
|  | Rice, white, long grain, boiled |
|  | Seeds of sunflower, linseed, and pumpkin, whole |
|  | Seeds, chia, dried |
|  | Sesame Butter or tahini |
|  | Swiss Chard |
|  | Tea, Herbal, Prepared |
|  | Tempeh or Tempe |
|  | Tomatoes, Red |
| Processed culinary ingredients (PCI) | Oil, olive |
|  | Oil, Soybean |
|  | Salt, table |
|  | Sugar, white granulated |
|  | Syrup, corn or sugar |
|  | Vegan Butter |
| Processed foods (PF) | Bread, French |
|  | Bread, wheat |
|  | Candy Bar, Peanut |
|  | Cereal, Granola, Prepared |
|  | Crackers, Rice |
|  | Crackers, water biscuits |
|  | Jelly |
|  | Pasta, made without egg, cooked |
|  | Tofu, firm, with calcium and magnesium chloride |
| Ultra-processed foods (UPF) | Acai berry drink |
|  | Almond milk |
|  | Bread, Mixed Grain |
|  | Cereal Bar, Rice and Wheat |
|  | Cheese, imitation |
|  | Cookie |
|  | Crackers, whole wheat |
|  | Pea protein supplements |
|  | Pizza |
|  | Potatoes or French Fries |
|  | Sauce, Soy (Shoyu) |
|  | Soda, cola |
|  | Soy protein - Supplements |
|  | Soymilk, Calcium Fortified |
|  | Textured Soy Protein |
|  | Vegan Nuggets, Baked |
|  | Vegan Protein Sausages |
|  | Veggie Burger, Unprepared |
|  | Yogurt, Tofu |
|  | Vegan Mayo |

**eTable 2.** Comparison of subsets with available and missing data across variables

| **Characteristic** | **Overall, N = 774** | **Subset with available data, N = 558** | **Subset with missing anthropometric data, N = 216** |
| --- | --- | --- | --- |
| Age | 29 (24, 35) | 29 (24, 37) | 28 (23, 33) |
| Sex |  |  |  |
| Female | 637 (82%) | 459 (82%) | 178 (82%) |
| Male | 137 (18%) | 99 (18%) | 38 (18%) |
| Body weight (kg) | 60 (54, 71) | 60 (54, 71) | - |
| Height (cm) | 165 (160, 170) | 165 (160, 170) | - |
| BMI (g/kg²) | 22.6 (20.3, 24.8) | 22.6 (20.3, 24.8) | - |
| Educational level |  |  |  |
| College education or technician, complete | 242 (31%) | 176 (32%) | 66 (31%) |
| Elementary school, completed | 2 (0.3%) | 1 (0.2%) | 1 (0.5%) |
| Elementary school, incomplete | 2 (0.3%) | 1 (0.2%) | 1 (0.5%) |
| High school, completed | 55 (7.1%) | 32 (5.7%) | 23 (11%) |
| High school, incomplete | 18 (2.3%) | 9 (1.6%) | 9 (4.2%) |
| Postgraduate | 280 (36%) | 219 (39%) | 61 (28%) |
| Undergoing college or technician education | 175 (23%) | 120 (22%) | 55 (25%) |
| Income |  |  |  |
| A class | 41 (5.3%) | 23 (4.1%) | 18 (8.3%) |
| B class | 296 (38%) | 222 (40%) | 74 (34%) |
| C class | 196 (25%) | 147 (26%) | 49 (23%) |
| D/E class | 241 (31%) | 166 (30%) | 75 (35%) |
| Smoking status | 68 (8.8%) | 45 (8.1%) | 23 (11%) |
| Alcohol consumption |  |  |  |
| No alcohol consumption | 324 (42%) | 236 (42%) | 88 (41%) |
| Once to twice a month | 147 (19%) | 108 (19%) | 39 (18%) |
| Twice to for times a month | 222 (29%) | 162 (29%) | 60 (28%) |
| Twice to three times per week | 72 (9.3%) | 46 (8.2%) | 26 (12%) |
| For or move times per week | 9 (1.2%) | 6 (1.1%) | 3 (1.4%) |
| Habitual physical exercise |  |  |  |
| Does not exercise | 142 (18%) | 108 (19%) | 34 (16%) |
| 1-2 hour/week | 156 (20%) | 111 (20%) | 45 (21%) |
| 7 hour/week or more | 124 (16%) | 91 (16%) | 33 (15%) |
| 5-6 hour/week | 176 (23%) | 122 (22%) | 54 (25%) |
| 3-4 hour/week | 176 (23%) | 126 (23%) | 50 (23%) |
| How long as a vegan |  |  |  |
| Less than one year | 110 (14%) | 72 (13%) | 38 (18%) |
| 1 to 2 years | 174 (22%) | 123 (22%) | 51 (24%) |
| 2 to 3 years | 153 (20%) | 111 (20%) | 42 (19%) |
| 3 to 4 years | 118 (15%) | 94 (17%) | 24 (11%) |
| 5 or more years | 219 (28%) | 158 (28%) | 61 (28%) |
| Supplement use | 592 (76%) | 430 (77%) | 162 (75%) |
| Kilocalories (kcal) | 1,782 (1,385, 2,227) | 1,777 (1,393, 2,257) | 1,805 (1,356, 2,225) |
| Protein (g) | 70 (48, 94) | 69 (48, 93) | 72 (50, 98) |
| Carbohydrate (g) | 268 (204, 346) | 266 (204, 345) | 270 (205, 348) |
| Fat (g) | 53 (37, 72) | 53 (37, 73) | 51 (36, 68) |
| Protein (g/kg) | 1.12 (0.79, 1.53) | 1.12 (0.79, 1.53) | - |
| Protein (% TEI) | 14.9 (12.3, 18.8) | 14.7 (12.0, 18.5) | 15.3 (12.8, 19.7) |
| Carbohydrate (% TEI) | 59 (52, 65) | 58 (52, 65) | 60 (54, 65) |
| Fat (% TEI) | 25 (19, 31) | 25 (19, 31) | 24 (18, 29) |
| Saturated fatty acid (g) | 10 (7, 14) | 10 (7, 15) | 9 (6, 14) |
| Monounsatured fatty acid (g) | 19 (13, 28) | 19 (13, 28) | 17 (13, 26) |
| Polyunsaturated fatty acid (g) | 16 (11, 22) | 16 (11, 22) | 16 (10, 22) |
| Trans-fatty acid (g) | 0.04 (0.01, 0.08) | 0.04 (0.01, 0.08) | 0.04 (0.02, 0.09) |
| Sodium (mg) | 2,376 (1,536, 3,233) | 2,392 (1,589, 3,234) | 2,281 (1,407, 3,218) |
| Potassium (mg) | 3,626 (2,716, 4,638) | 3,614 (2,692, 4,629) | 3,762 (2,847, 4,663) |
| Vitamin A (retinol) (mcg) | 869 (403, 1,642) | 870 (403, 1,685) | 867 (402, 1,511) |
| Vitamin A (IU) | 8,419 (3,922, 16,119) | 8,419 (3,987, 16,487) | 8,475 (3,827, 14,767) |
| Beta carotene (µg) | 4,317 (1,801, 8,049) | 4,412 (1,805, 8,399) | 4,049 (1,786, 7,286) |
| Alpha carotene (µg) | 710 (96, 2,102) | 712 (95, 2,139) | 700 (98, 2,073) |
| Lutein (µg) | 1,979 (942, 4,227) | 1,998 (959, 4,340) | 1,967 (933, 3,900) |
| Cryptoxanthin beta (µg) | 129 (7, 729) | 151 (7, 709) | 79 (8, 738) |
| Lycopene (µg) | 1,831 (8, 4,566) | 1,894 (12, 4,570) | 1,686 (2, 4,201) |
| Vitamin C (mg) | 157 (81, 248) | 155 (81, 248) | 161 (80, 247) |
| Calcium (mg) | 571 (371, 841) | 582 (370, 850) | 544 (376, 785) |
| Iron (mg) | 17 (13, 23) | 17 (13, 23) | 17 (13, 23) |
| Vitamin D (µg) | 0.00 (0.00, 2.20) | 0.00 (0.00, 2.13) | 0.02 (0.00, 2.22) |
| Vitamin D (IU) | 0 (0, 86) | 0 (0, 86) | 0 (0, 88) |
| Vitamin E (mg) | 0.00 (0.00, 0.45) | 0.00 (0.00, 0.47) | 0.00 (0.00, 0.38) |
| Vitamin E (IU) | 0.0 (0.0, 0.7) | 0.0 (0.0, 0.7) | 0.0 (0.0, 0.6) |
| Alpha-tocopherol (vitamin E) (mg) | 9 (6, 14) | 9 (7, 14) | 9 (6, 14) |
| Thiamin (mg) | 1.69 (1.25, 2.32) | 1.68 (1.26, 2.28) | 1.74 (1.23, 2.44) |
| Riboflavin (mg) | 1.09 (0.82, 1.47) | 1.08 (0.81, 1.47) | 1.10 (0.82, 1.45) |
| Niacin (mg) | 14 (10, 19) | 14 (10, 19) | 15 (11, 19) |
| Pyridoxine (mg) | 1.82 (1.35, 2.51) | 1.81 (1.35, 2.50) | 1.83 (1.33, 2.52) |
| Folate (mg) | 703 (506, 941) | 701 (515, 941) | 704 (494, 941) |
| Cobalamin (mg) | 0.05 (0.00, 1.78) | 0.05 (0.00, 1.76) | 0.11 (0.00, 2.00) |
| Biotin (mg) | 14 (8, 24) | 14 (8, 24) | 13 (8, 24) |
| Pantothenic (mg) | 4.12 (3.20, 5.73) | 4.11 (3.16, 5.72) | 4.22 (3.34, 5.74) |
| Vitamin K (µg) | 122 (72, 215) | 121 (72, 212) | 131 (68, 216) |
| Phosphorus (mg) | 1,130 (803, 1,509) | 1,123 (808, 1,503) | 1,142 (797, 1,542) |
| Magnesium (mg) | 442 (329, 594) | 440 (332, 602) | 444 (321, 587) |
| Zinc (mg) | 8.2 (5.9, 10.8) | 8.2 (5.9, 10.8) | 8.3 (5.8, 10.8) |
| Copper (mg) | 1.95 (1.42, 2.65) | 1.93 (1.46, 2.65) | 1.99 (1.35, 2.64) |
| Manganese (mg) | 6.0 (4.2, 8.5) | 6.1 (4.2, 8.7) | 6.0 (4.3, 8.3) |
| Selenium (µg) | 61 (44, 84) | 60 (43, 83) | 62 (46, 84) |
| Fluoride (µg) | 198 (74, 377) | 197 (71, 383) | 200 (94, 342) |
| Chromium (mg) | 0.05 (0.03, 0.09) | 0.05 (0.03, 0.09) | 0.05 (0.03, 0.09) |
| Molybdenum (mg) | 5 (1, 17) | 5 (1, 17) | 5 (1, 15) |
| Choline (mg) | 199 (145, 263) | 199 (143, 262) | 199 (147, 264) |
| Dietary fiber (g) | 44 (31, 61) | 44 (31, 60) | 43 (30, 63) |
| Results are presented as median (interquartile range) for continuous variables and n (%) for categorical variables. BMI, body mass index. | | | |

**eTable 3.** Micronutrients intake

| **Characteristic** | **Overall**, N = 774 | **Female**, N = 637 | **Male**, N = 137 |
| --- | --- | --- | --- |
| **24-hour food recall** |  |  |  |
| Sodium (mg) | 2,376 (1,536, 3,233) | 2,282 (1,486, 3,106) | 2,871 (1,833, 4,091) |
| Potassium (mg) | 3,626 (2,716, 4,638) | 3,452 (2,628, 4,418) | 4,389 (3,308, 6,055) |
| Vitamin A (retinol) (mcg) | 869 (403, 1,642) | 842 (412, 1,602) | 1,040 (341, 1,880) |
| Vitamin C (mg) | 157 (81, 248) | 154 (79, 245) | 172 (87, 270) |
| Calcium (mg) | 571 (371, 841) | 565 (357, 827) | 598 (407, 881) |
| Iron (mg) | 17 (13, 23) | 17 (12, 22) | 21 (15, 28) |
| Alpha-tocopherol (vitamin E) (mg) | 9 (6, 14) | 9 (6, 13) | 12 (8, 17) |
| Thiamin (mg) | 1.69 (1.25, 2.32) | 1.62 (1.21, 2.15) | 2.24 (1.61, 2.89) |
| Riboflavin (mg) | 1.09 (0.82, 1.47) | 1.06 (0.77, 1.43) | 1.28 (1.01, 1.71) |
| Niacin (mg) | 14 (10, 19) | 14 (10, 18) | 18 (14, 23) |
| Pyridoxine (mg) | 1.82 (1.35, 2.51) | 1.73 (1.29, 2.35) | 2.38 (1.66, 3.22) |
| Folate (mg) | 703 (506, 941) | 670 (487, 887) | 875 (624, 1,133) |
| Cobalamin (mg) | 0.05 (0.00, 1.78) | 0.06 (0.00, 1.76) | 0.05 (0.00, 2.04) |
| Biotin (mg) | 14 (8, 24) | 13 (7, 23) | 18 (9, 33) |
| Pantothenic acid (mg) | 4.12 (3.20, 5.73) | 3.95 (3.07, 5.40) | 5.39 (3.64, 6.65) |
| Vitamin K (µg) | 122 (72, 215) | 118 (70, 214) | 139 (82, 216) |
| Phosphorus (mg) | 1,130 (803, 1,509) | 1,092 (791, 1,440) | 1,367 (933, 1,869) |
| Magnesium (mg) | 442 (329, 594) | 430 (315, 567) | 529 (381, 763) |
| Zinc (mg) | 8.2 (5.9, 10.8) | 7.9 (5.7, 10.2) | 10.4 (6.9, 13.7) |
| Copper (mcg) | 1950 (1420, 2650) | 1.870 (1380, 2540) | 2380 (1660, 3330) |
| Manganese (mg) | 6.0 (4.2, 8.5) | 5.7 (4.0, 7.9) | 8.0 (5.0, 10.8) |
| Selenium (µg) | 61 (44, 84) | 59 (43, 78) | 79 (50, 116) |

Results are presented as median (interquartile range). Micronutrient intake reported herein does not consider the contribution from isolated micronutrient supplementation.

**eTable 4.** Caloric contribution and protein intake according to Nova food processing category and amino acid intake (considering textured soy protein as UMPF)

| **Food processing category** | **Overall**, N = 774 | **Female**, N = 637 | **Male**, N = 137 | **Metropolitan population reference value** |
| --- | --- | --- | --- | --- |
| *Caloric contribution* |  |  |  |  |
| UMPF (% TEI) | 68.6 (67.6, 70) | 68.7 (67.6, 70.3) | 68.2 (63.5, 70.8) | 44.9 |
| PCI (% TEI) | 8.3 (7.6, 8.8) | 8.4 (7.8, 9) | 8.1 (6.6, 9.4) | 19.4 |
| PF (% TEI) | 6.2 (5.1, 6.8) | 6.6 (5.7, 7.5) | 3.9 (0.6, 6.7) | 12.1 |
| UPF (% TEI) | 10.7 (9.4, 11.7) | 10.5 (9, 11.5) | 11 (8.5, 13.6) | 23.7 |
| *Protein intake according to processing* |  |  |  |  |
| UMPF (% total protein intake) | 71.0 (69.2, 72.6) | 71.2 (69.3, 73.6) | 70.7 (66.3, 74) | **-** |
| PF (% total protein intake) | 7.4 (6.1, 8.7) | 7.8 (6.4, 9.2) | 5.2 (2.5, 9.3) | **-** |
| UPF (% total protein intake) | 14.3 (12.2, 16) | 13.9 (11.9, 16.1) | 17.7 (13.3, 22) | **-** |

Results are presented as median (95% confidence interval). Confidence intervals show the estimated range containing the true population median for each variable, with 95% confidence. **UMPF:** unprocessed and minimally processed foods; **PCI:** processed culinary ingredient; **PF:** processed foods; **UPF:** ultra-processed foods.

**eTable 5.** Logistic regression model coefficients (complete case analysis, n=558)

| **Independent variable** | **Response** | **Odds ratio (95% CI)** | | **P-value** |
| --- | --- | --- | --- | --- |
| Isolated protein supplement consumer | No | 1.00 (reference) | | - |
|  | Yes | 0.05 (0.01, 0.12) | | <0.0001 |
| Textured soy protein consumer | No | 1.00 (reference) | | - |
|  | Yes | 0.31 (0.16, 0.58) | | <0.0001 |
| **Independent variable** | **Quartile** | **Value** | **Odds ratio (95% CI)** | **P-value** |
| Total kilocalories intake | 1^st^ quartile | 1392 Kcal | 1.00 (reference) | - |
|  | 2^nd^ quartile | 1776 Kcal | 0.17 (0.09, 0.30) | <0.0001 |
|  | 3^rd^ quartile | 2257 Kcal | 0.05 (0.02, 0.09) | <0.0001 |
|  | 4^th^ quartile | > 2257 Kcal | 0.01 (0.003, 0.03) | <0.0001 |
| UMPF intake | 1^st^ quartile | 54.5 % TEI | 1.00 (reference) | - |
|  | 2^nd^ quartile | 67.2 % TEI | 0.68 (0.38, 1.21) | 0.20 |
|  | 3^rd^ quartile | 76.7 % TEI | 0.99 (0.56, 1.76) | 0.98 |
|  | 4^th^ quartile | > 76.7 % TEI | 1.19 (0.68, 2.10) | 0.53 |
| UPF intake | 1^st^ quartile | 3.7 % TEI | 1.00 (reference) | - |
|  | 2^nd^ quartile | 12.3 % TEI | 0.31 (0.17, 0.54) | <0.0001 |
|  | 3^rd^ quartile | 23.0 % TEI | 0.27 (0.15, 0.49) | <0.0001 |
|  | 4^th^ quartile | > 23.0 % TEI | 0.29 (0.16, 0.50) | <0.0001 |
| Unprocessed protein intake | 1^st^ quartile | 46.0 % TEI | 1.00 (reference) | - |
|  | 2^nd^ quartile | 62.6 % TEI | 3.33 (1.72, 6.68) | <0.001 |
|  | 3^rd^ quartile | 81.2 % TEI | 2.89 (1.48, 5.84) | 0.002 |
|  | 4^th^ quartile | > 81.2 % TEI | 5.71 (3.02, 11.28) | <0.0001 |
| Ultra-processed protein intake | 1^st^ quartile | 5.1 % TEI | 1.00 (reference) | - |
|  | 2^nd^ quartile | 21.9 % TEI | 0.61 (0.36, 1.04) | 0.07 |
|  | 3^rd^ quartile | 41.1 % TEI | 0.22 (0.12, 0.40) | <0.0001 |
|  | 4^th^ quartile | > 41.1 % TEI | 0.12 (0.06, 0.24) | <0.0001 |

**eTable 6.** Logistic regression model coefficients (imputed dataset, n=774)

| **Independent variable** | **Response** | **Odds ratio (95% CI)** | | **P-value** |
| --- | --- | --- | --- | --- |
| Isolated protein supplement consumer | No | 1.00 (reference) | | - |
|  | Yes | 0.04 (0.01, 0.09) | | <0.0001 |
| Textured soy protein consumer | No | 1.00 (reference) | | - |
|  | Yes | 0.24 (0.13, 0.43) | | <0.0001 |
| **Independent variable** | **Quartile** | **Value** | **Odds ratio (95% CI)** | **P-value** |
| Total kilocalories intake | 1^st^ quartile | 1392 Kcal | 1.00 (reference) | - |
|  | 2^nd^ quartile | 1776 Kcal | 0.14 (0.08, 0.23) | <0.0001 |
|  | 3^rd^ quartile | 2257 Kcal | 0.04 (0.02, 0.07) | <0.0001 |
|  | 4^th^ quartile | > 2257 Kcal | 0.006 (0.002, 0.017) | <0.0001 |
| UMPF intake | 1^st^ quartile | 54.5 % TEI | 1.00 (reference) | - |
|  | 2^nd^ quartile | 67.2 % TEI | 0.63 (0.38, 1.03) | 0.07 |
|  | 3^rd^ quartile | 76.7 % TEI | 0.89 (0.55, 1.45) | 0.64 |
|  | 4^th^ quartile | > 76.7 % TEI | 1.02 (0.63, 1.64) | 0.93 |
| UPF intake | 1^st^ quartile | 3.7 % TEI | 1.00 (reference) | - |
|  | 2^nd^ quartile | 12.3 % TEI | 0.33 (0.20, 0.54) | <0.0001 |
|  | 3^rd^ quartile | 23.0 % TEI | 0.32 (0.20, 0.52) | <0.0001 |
|  | 4^th^ quartile | > 23.0 % TEI | 0.32 (0.19, 0.51) | <0.0001 |
| Unprocessed protein intake | 1^st^ quartile | 46.0 % TEI | 1.00 (reference) | - |
|  | 2^nd^ quartile | 62.6 % TEI | 2.69 (1.55, 4.79) | <0.001 |
|  | 3^rd^ quartile | 81.2 % TEI | 2.44 (1.39, 4.38) | 0.002 |
|  | 4^th^ quartile | > 81.2 % TEI | 5.05 (2.96, 8.88) | <0.0001 |
| Ultra-processed protein intake | 1^st^ quartile | 5.1 % TEI | 1.00 (reference) | - |
|  | 2^nd^ quartile | 21.9 % TEI | 0.61 (0.39, 0.95) | 0.03 |
|  | 3^rd^ quartile | 41.1 % TEI | 0.28 (0.17, 0.46) | <0.0001 |
|  | 4^th^ quartile | > 41.1 % TEI | 0.13 (0.07, 0.22) | <0.0001 |

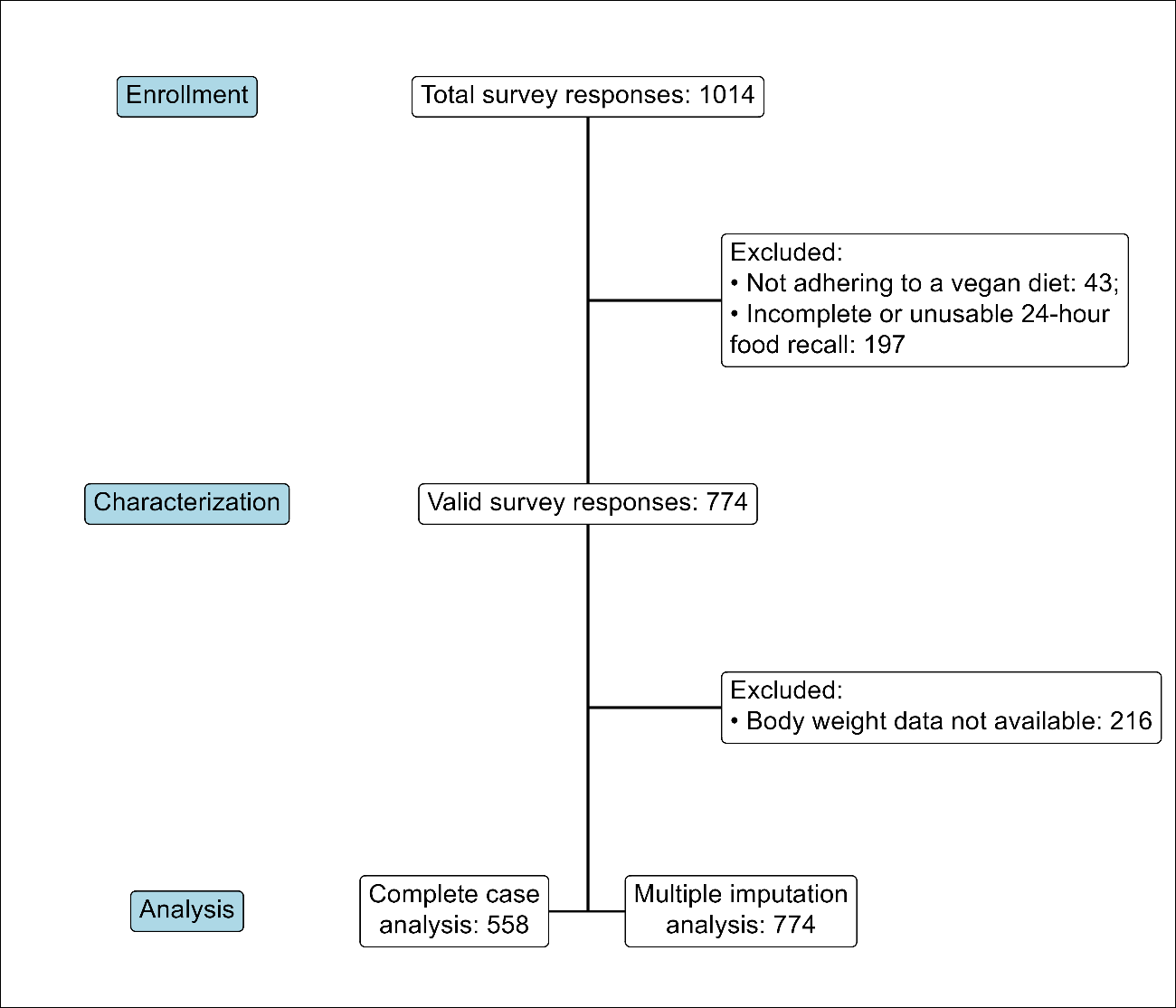

**eFigure 1.** Study flowchart

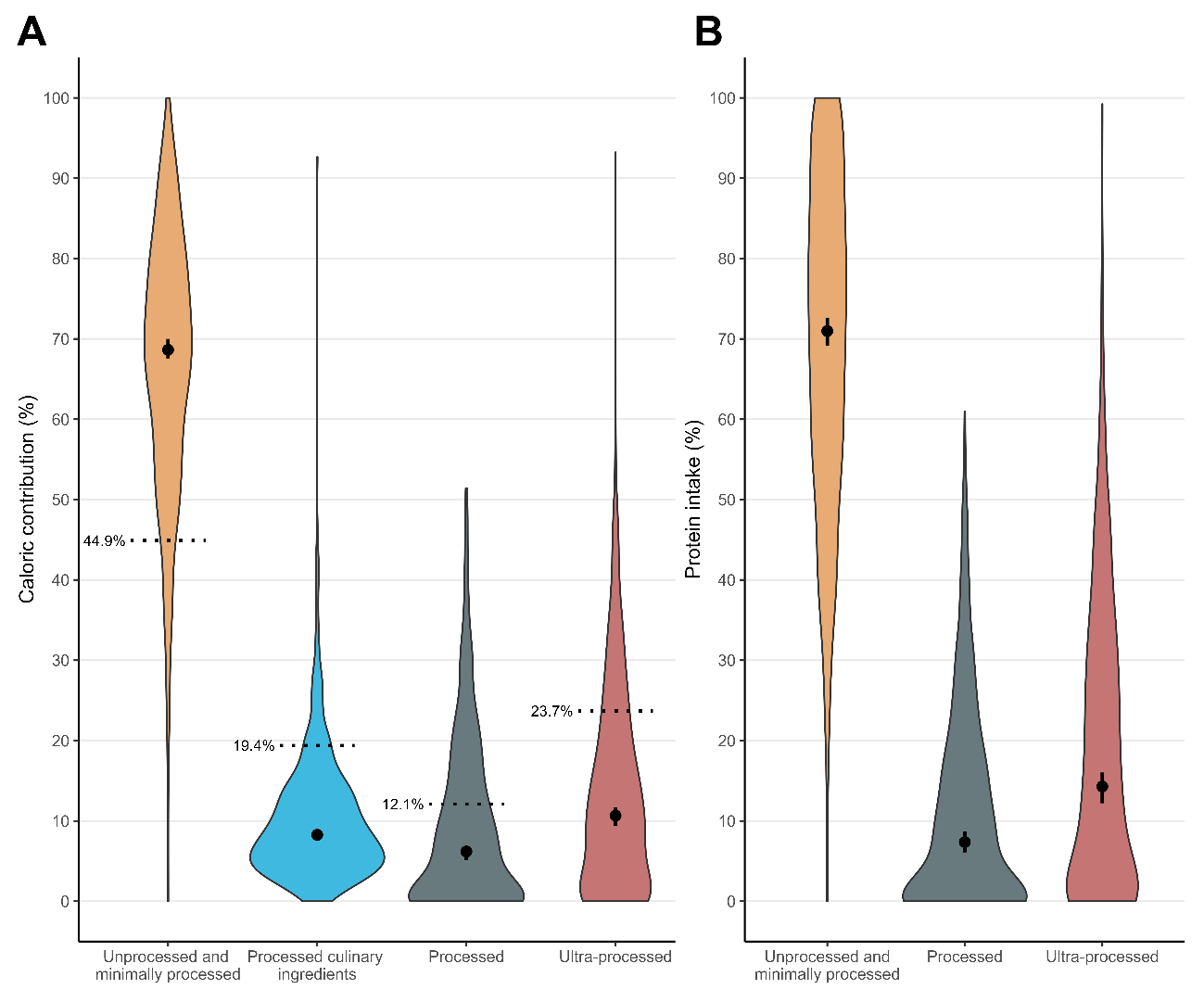

**eFigure 2.** Caloric and protein intake according to Nova food processing categories (considering textured soy protein as UMPF). Panel A: Violin plots showing the distribution of caloric contribution of each food processing category. Dashed line show reference values from the Brazilian population living in metropolitan areas. Panel B: protein intake contribution of each food processing category. Dots are medians accompanied by 95% confidence intervals.

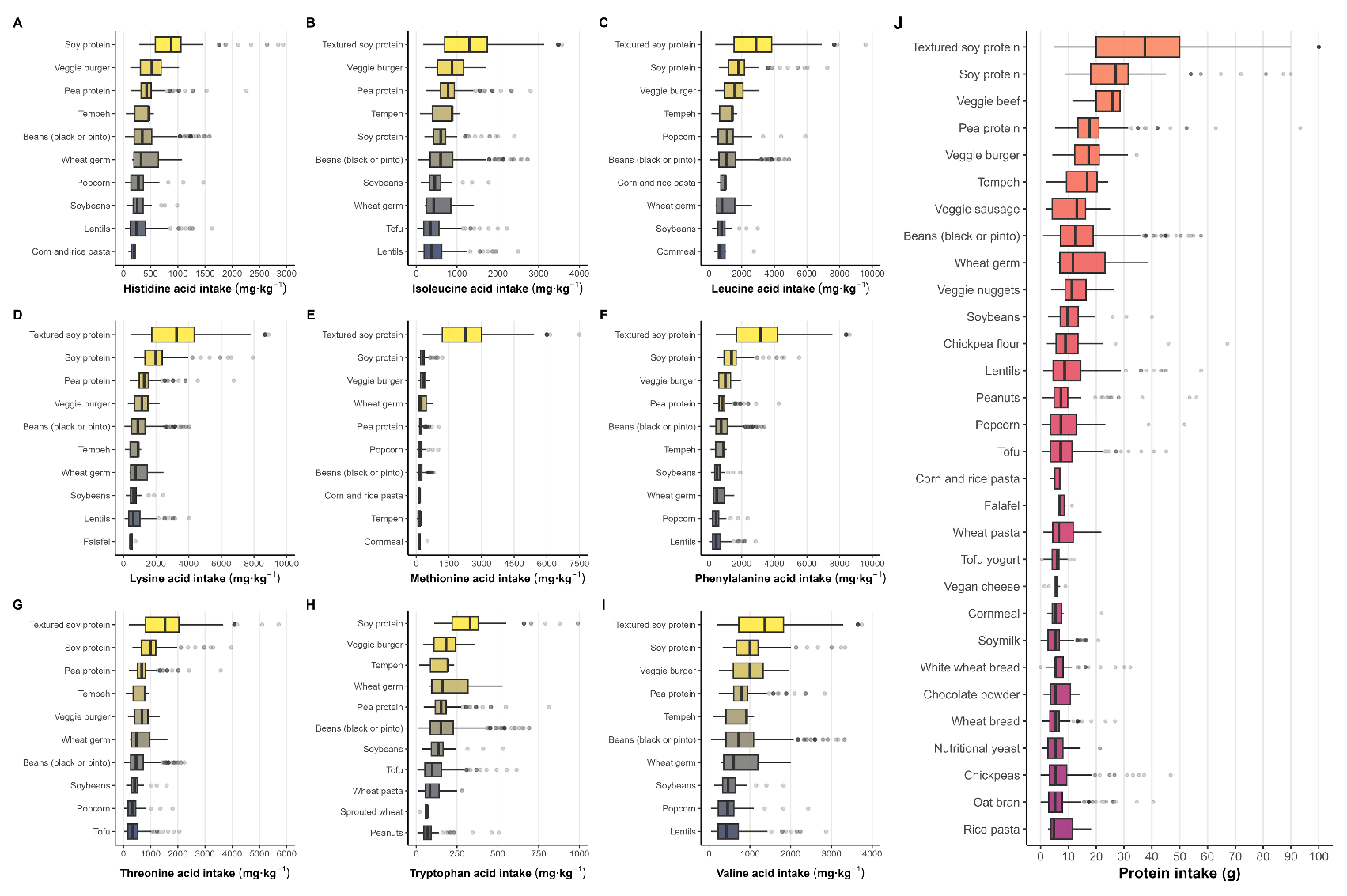

**eFigure 3.** Main food sources of protein and essential amino acids.

Caption: Boxplots showing protein and essential amino acid intake contribution for the top 30 and top 10 foods, respectively. Panel A: Histidine intake. Panel B: Isoleucine intake. Panel C: Leucine intake. Panel D: Lysine intake. Panel E: Methionine intake. Panel F: Phenylalanine intake. Panel G: Threonine intake. Panel H: Tryptophane intake. Panel I: Valine intake. Panel J: Protein intake.

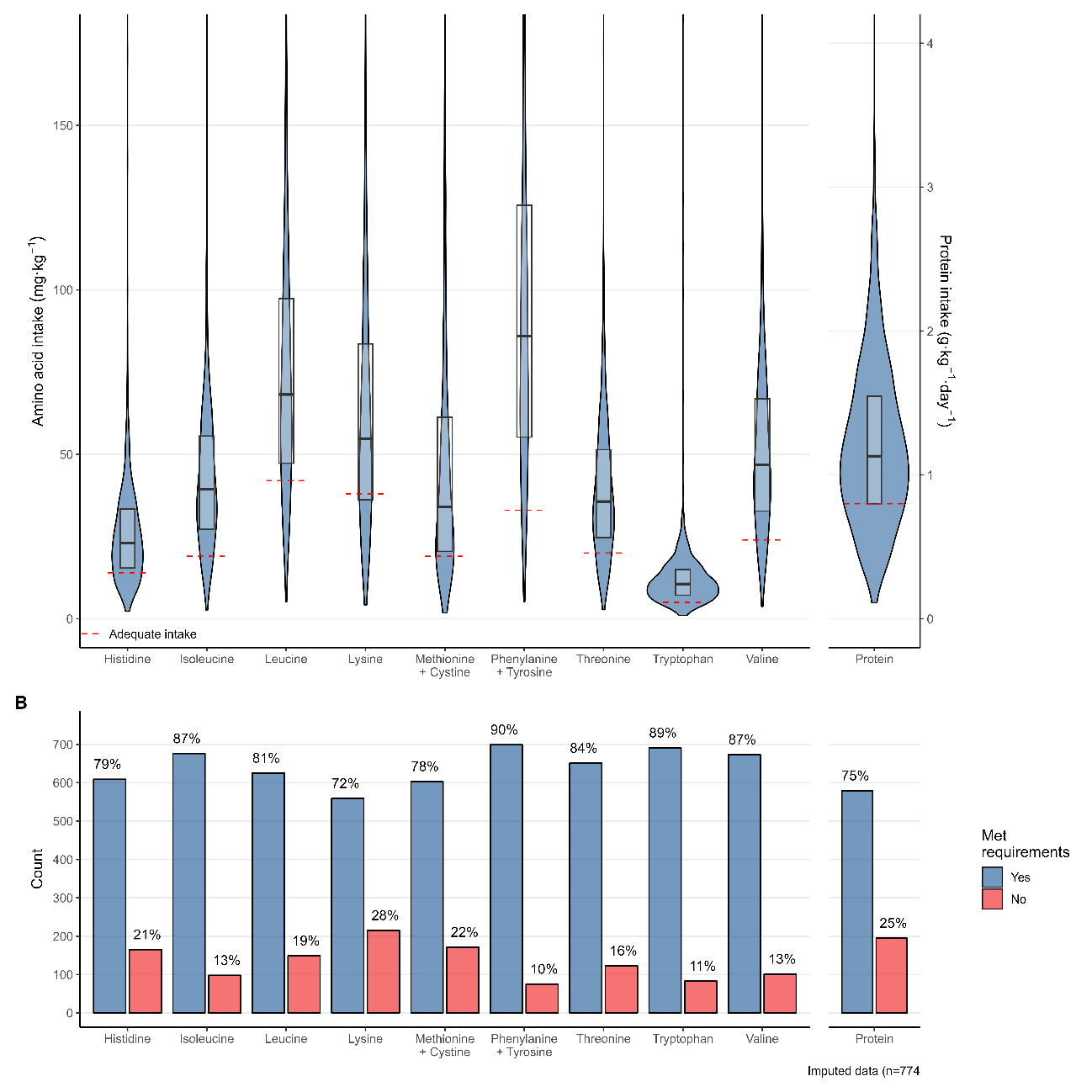
**eFigure 4.** Essential amino acids intake and proportion of individuals attaining RDAs. Panel A: Violin plots showing the distribution of essential amino acids intake relative to body mass, with the red dashed line indicating the respective RDA and boxplots showing median values and interquartile range. Panel B: Count and proportion of individuals meeting RDA for essential amino acids intake (second plot). Missing body weight values (n = 216) were multiply imputed, totalling n = 774.
